# Clinical Evaluation of the GeneXpert^®^ Xpert^®^ Xpress SARS-CoV-2/Flu/RSV *PLUS* Combination Test

**DOI:** 10.1101/2022.11.07.22281957

**Authors:** Grant Johnson, Branden S.J. Gregorchuk, Arek Zubrzycki, Kurt Kolsun, Adrienne F.A. Meyers, Paul A. Sandstrom, Michael G. Becker

## Abstract

The GeneXpert^®^ Xpert^®^ Xpress SARS-CoV-2/Flu/RSV *PLUS* combination test (*PLUS* Assay) received Health Canada approval in January 2022. The *PLUS* Assay is similar to the SARS-CoV-2/Flu/RSV combination test, with modifications to improve assay robustness against circulating and emerging variants. The performance characteristics of the *PLUS* Assay were assessed at the Lakeridge Health Oshawa Hospital Centre and the National Microbiology Laboratory of Canada. The *PLUS* Assay was directly compared to the SARS-CoV-2/Flu/RSV combination test using SARS-CoV-2 culture from five variants and remnant clinical specimens collected across the COVID-19 pandemic. This included 50 clinical specimens negative for all pathogens, 110 clinical specimens positive for SARS-CoV-2, Influenza A, Influenza B, RSVA, and/or RSVB and an additional 11 mixed samples to screen for target interactions. The *PLUS* Assay showed a high percent agreement with the widely used SARS-CoV-2/Flu/RSV combination test. Based on these findings, the *PLUS* Assay and the Xpert SARS-CoV-2/Flu/RSV combination test results are largely consistent with no observed difference in sensitivity, specificity, or time to result when challenged with various SARS-CoV-2 variants. The reported Ct values provided by the new *PLUS* Assay was also unchanged, with the exception of a possible 1-2 decrease reported Ct for RSVA across a limited sample size.

## Introduction

The coronavirus disease 2019 (COVID-19) pandemic has lead to an enormous demand for diagnostic testing. To this end, numerous antigen and molecular SARS-CoV-2 rapid tests have been approved by the United States Food and Drug Administration and Health Canada, including the Cepheid GeneXpert® Xpert® Xpress SARS-CoV-2/Flu/RSV Combination Test. The GeneXpert and its associated COVID-19 cartridges perform a rapid, fully-automated, and self-contained multiplex RT-qPCR tests with run times of 45 minutes or less (Cepheid Package Insert, 2021). The Xpert SARS-CoV-2/Flu/RSV assay targets two SARS-CoV-2 genomic regions, the envelope (E) and the nucleocapsid (N2) returning a cycle threshold (Ct) value if the target is detected as well as its qualitative result interpretation within 45 amplification cycles. The results for the SARS-CoV-2/Flu/RSV combination test reports the fluorescence values for both the E and N2 targets of SARS-CoV-2 as a single result as they occupy the same fluorescence channel (Johnson *et al*., 2021). Additionally, the Xpert SARS-CoV-2/Flu/RSV combination test is able to detect influenza A, influenza B as well as respiratory syncytial virus (RSV), and will report their respective Ct values.

As SARS-CoV-2 variants of concern continue to emerge worldwide, Cepheid developed a new formulation of their Xpert SARS-CoV-2/Flu/RSV combination test, the Xpert SARS-CoV-2/Flu/RSV *PLUS* combination test (referred to hereafter as the *PLUS* Combination Test). The *PLUS* Combination Test has an additional SARS-CoV-2 target, RNA-Dependent RNA Polymerase (RdRp), to help improve assay robustness against novel SARS-CoV-2 variants and redesigned N2 oligos for better coverage of nucleocapsid mutations in circulation that have been shown to cause N gene detection dropout using GeneXpert SARS-CoV-2 assays (Foster *et al*., 2022; Isabel *et al*., 2022). RdRp (also known as nsp-12) is the catalytic subunit of multi-subunit polymerase complex which, along with nsp-7 and nsp-8, forms the minimal core components for SARS-CoV-2 RNA synthesis (Peng *et al*., 2020). To date, there has been no identified RdRp modifications, with this region being considered relatively stable target due to its conserved function (te Velthuis *et al*., 2009; Pachetti *et al*., 2020; Peng *et al*., 2020). RdRp is measured on the same fluorescence channel as targets E and N2, and thus a single Ct value is reported for SARS-CoV-2.

As a rapid near-point-of-care device, the GeneXpert system is currently used as a testing option to improve turnaround times in major health centres, and is used heavily in critical care settings (Jokela *et al*., 2020; Gotham *et al*., 2021). Additionally, the GeneXpert has been a reliable tool for decentralized testing in remote and isolated Canadian communities (Respiratory Virus Infections Working Group, 2020) where access to large, well-equipped laboratories is not possible (Berry *et al*., 2020; Jokela *et al*., 2020; Gotham *et al*., 2021; Yau *et al*., 2021; Rong *et al*., 2022). It has also been used internationally by the World Health Organization in developing countries during their response to the COVID-19 pandemic (Rakotosamimanana *et al*., 2020; World Health Organization, 2020).

The individual Xpert Xpress SARS-CoV-2 and Xpert Xpress Flu/RSV assays have demonstrated high sensitivity in numerous analytical and clinical studies with company reported sensitivity of 100% (n=35) at 250 copies (cp)/mL for its SARS-CoV-2 assay; independent studies have reported a limit of detection (LOD) ranging from 8.3 – 60 cp/mL (3–4). Indeed, the assay has shown excellent agreement with the Roche Cobas 6800 system (Broder *et al*., 2020; Goldenberger *et al*., 2020; Lieberman *et al*., 2020; Moran *et al*., 2020; Smithgall *et al*., 2020; Tham *et al*., 2021), the Hologic Panther Fusion (Hogan *et al*., 2020), as well as laboratory-developed RT-qPCR tests (Lieberman *et al*., 2020; Wolters *et al*., 2020). A recent systematic review of the Xpert Xpress assay indicated an overall specificity and sensitivity of 97% based on a selection of 11 studies (Goldenberger *et al*., 2020). As the GeneXpert is present a plethora of settings (including clinics, hospitals, health centres, nursing stations), an independent evaluation of the recently approved *PLUS* Combination Test ahead of the approaching flu season is essential.

In our study, remnant clinical samples of SARS-CoV-2, influenza A (H3 and H1 2009), influenza B, RSVA, and RSVB were used to evaluate the *PLUS* Combination Test and compare it to the SARS-CoV-2/Flu/RSV Combination Test (Johnson *et al*., 2022). This evaluation was performed in two independent laboratories, the National Microbiology Laboratory (NML; Winnipeg, Canada) and Lakeridge Health Oshawa Hospital (Oshawa, Canada). The SARS-CoV-2 samples used in this study were collected across multiple waves of the pandemic, and included SARS-CoV-2 Alpha, Delta, Omicron BA.1, and Omicron BA.2 variants. Additionally, inactivated SARS-CoV-2 culture from five variants was tested on both assays. No difference was observed between the assays concerning sensitivity, reported Ct value, run time, or specificity.

## Materials and Methods

### Clinical Specimens

Remnant universal transport media (UTM) from nasopharyngeal or nasal clinical swabs collected at the Cadham Provincial Laboratory (Winnipeg, Canada), Lakeridge Health Oshawa Hospital (Oshawa, Canada), and the Public Health Ontario Laboratory (Toronto, Canada) were used. Study samples included 50 clinical specimens negative for SARS-CoV-2, Influenza A, Influenza B, and RSV, with an additional 99 clinical positive specimens containing any one or more of the following pathogens: SARS-CoV-2, Influenza A, Influenza B, or RSV. Fifty-nine of the study samples (50 negatives, 7 COVID-19 positive, 2 Influenza positive) were obtained from prospective sampling of patients presenting with COVID-19 symptoms. The remainder of the study samples were previously characterized by laboratory-developed RT-qPCR tests (i.e. the ResPlex Assay or the Cepheid GeneXpert with the SARS-CoV-2/Flu/RSV Combination Test; Supplemental Data 1). Selected samples were stratified to cover a wide range of Ct values, from approximately 15-40. All samples tested at the National Microbiology Laboratory of Canada each RSV sample were typed as RSVA or RSVB, which is indicated in Supplemental Data 1. In total, this study included 7 RSVA samples, 9 RSVB samples, and 10 RSV samples that were not typed.

To simulate patient coinfection with multiple viruses, remnant transport media from an additional 26 clinical specimens were mixed in various combinations to create 11 contrived samples, each containing two or three different respiratory viral pathogens, and tested using both Xpert assays (Supplemental Data 1). All samples used in this study were research ethics board-exempt, anonymized, diagnostic samples used for assay validation. To determine overall test agreement, 300 μL of transport media from nasal or nasopharyngeal swabs was tested in parallel with the SARS-CoV-2/Flu/RSV combination test and the *PLUS* Combination Test..

### Gamma-irradiated SARS-CoV-2 culture

High-titre inactivated SARS-CoV-2 culture of the Alpha (B.1.1.7), Delta (B.1.617.2), Omicron BA.1 (BA.1), Omicron BA.2 (BA.2), and wild type (WA-1) variants was provided by the Special Pathogens Program of the National Microbiology Laboratory Branch (NMLB; Winnipeg, MB). SARS-CoV-2 variants were propagated in Vero cells in Minimal Essential Medium and clarified by low speed centrifugation. The viral supernatant was inactivated via gamma-irradiation using a Gammacell 220 Cobalt-60 irradiator with a total exposure of three Mrad of radiation. The reported stock concentrations of the viral variant preparations were 2.1×10^6^ PFU/mL (Alpha), 2.3×10^5^ PFU/mL (Delta), 2.1×10^5^ PFU/mL (Omicron BA1), 1.9×10^5^ PFU/mL (Omicron BA.2), and 1.2×10^6^ PFU/mL (wild type). Inactivated culture fluid was serially diluted in UTM and Ct values were determined using the GeneXpert® Xpert® Xpress SARS-CoV-2 assay. Dilutions were performed to obtain Ct values of approximately 33 and 36, approaching the limit of detection of the GeneXpert assay. Ct values were converted into cp/mL using a standard curve (Becker *et al*., 2020). Characterization of dilutions are summarized in Table 1. 300 μL of the diluted culture fluid was tested in duplicate using both the SARS-CoV-2/Flu/RSV combination test and the *PLUS* Combination Test.

**Table 1:** Detection of five SARS-CoV-2 Variants. Materials tested were cultured SARS-CoV-2, inactivated by gamma-irradiation. SARS-CoV-2 concentrations were adjusted to target Ct values of 33 and 36, followed by testing with the Xpert SARS-CoV-2/Flu/RSV *PLUS* Combination Test or Xpert SARS-CoV-2/Flu/RSV Combination Test. Each dilution was tested in duplicate (indicated as Replicate 1 [Rep1] and Replicate 2 [Rep2] below).

|  | SARS-CoV-2 Variant<br>(inactivated culture) | SARS-CoV-2<br>/Flu/RSV |  | SARS-CoV-2<br>/Flu/RSV *PLUS* |  |
| --- | --- | --- | --- | --- | --- |
|  |  | Rep1 | Rep2 | Rep1 | Rep2 |
| Ct = 33 | Wild Type WA-1 | 30.2 | 32.2 | 32.9 | 32.8 |
|  | Alpha B.1.1.7 | 32.2 | 32.1 | 33.4 | 33.2 |
|  | Delta B.1.617.2 | 33.7 | 33.8 | 34.2 | 34.8 |
|  | Omicron BA.1.1 | 33.7 | 32.3 | 33.2 | 32.7 |
|  | Omicron BA.2 | 33.7 | 33.4 | 33.0 | 32.4 |
| Ct = 36 | Wild Type WA-1 | 35.8 | 35.5 | 36.1 | 36.0 |
|  | Alpha B.1.1.7 | 35.4 | 35.7 | 35.6 | 36.1 |
|  | Delta B.1.617.2 | 36.4 | 36.4 | 37.3 | 37.5 |
|  | Omicron BA.1.1 | 36.4 | 37.1 | 35.8 | 35.9 |
|  | Omicron BA.2 | 36.2 | 37.6 | 36.0 | 35.3 |

Note: Ct: Cycle threshold

## Results

Inactivated gamma-irradiated SARS-CoV-2 culture of five isolates (Wild-Type (WA-1), Alpha (B.1.1.7), Delta (B.1.617.2), Omicron (BA.1), and Omicron (BA.2)) were used to investigate the performance of the *PLUS* Combination Test against current or previous circulating variants. A panel of five cultured SARS-CoV-2 isolates was tested with the SARS-CoV-2/Flu/RSV combination test and the *PLUS* Combination Test (Table 1). All variants were detected with both assays at all dilutions and replicates. Additionally, SARS-CoV-2 positive and SARS-CoV-2 negative clinical samples were tested with both the *PLUS* Combination Test and SARS-CoV-2/Flu/RSV combination test (Table 2). The Ct values of SARS-CoV-2 positive samples ranged from 15.6 to 43.5 (Fig. 1; Supplemental Data 1). There was 100% agreement between both assays, with 57 positives and 103 negatives detected (Table 2). There was a high level of correlation between the SARS-CoV-2 Ct values reported with both of the assays (Figure 1; R^2^ = 0.973), with an average standard deviation of 0.49 Ct values between the reported results (Supplemental Data 1).

**Table 2:** Concordance of the Xpert SARS-CoV-2/Flu/RSV *PLUS* Combination Test with the Xpert SARS-CoV-2/Flu/RSV PLUS Combination Test for the detection of SARS-CoV-2.

| Result with<br>Xpert SARS-CoV-2/Flu/RSV | Result with Xpert SARS-CoV-2/Flu/RSV *PLUS* |  |
| --- | --- | --- |
|  | SARS-CoV-2 |  |
|  | Positive | Negative |
| Positive | 57 | 0 |
| Negative | 0 | 103 |

**Figure 1:**
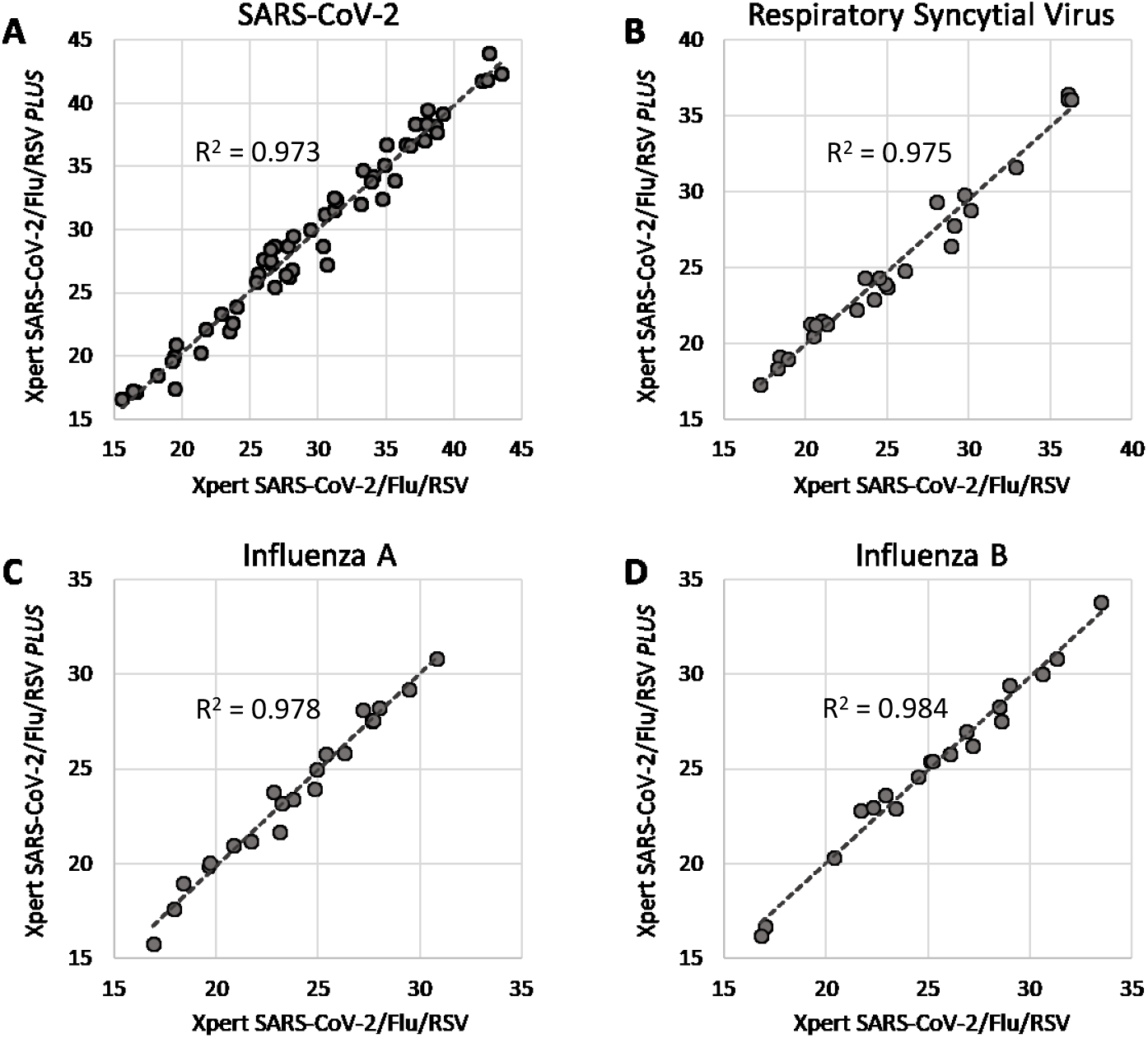
Ct value comparisons between the Xpert SARS-CoV-2/Flu/RSV Combination Test and SARS-CoV-2/Flu/RSV *PLUS* Combination Test. Results shown for: A) SARS-CoV-2; B) Respiratory Syncytial Virus; C) Influenza A; and D) Influenza B. Correlation (R^2^) is calculated for each target.

The above process was repeated for clinical samples containing Influenza A, Influenza B, and RSV. Similarly, results were highly consistent between the *PLUS* Combination Test and SARS-CoV-2/Influenza/RSV combination test, with overall percent agreements of 100%, 99.4%, and 100% for influenza A, influenza B, and RSV, respectively (Table 3). Regardless of RSV subtype, all RSV positive clinical samples were detected using both assays (Table 3; Supplemental Data 1). Likewise, high correlations were observed between the Ct values reported by both Xpert assays for influenza A (Figure 1; R^2^ = 0.978), influenza B (R^2^ = 0.984), and RSV (R^2^ = 0.975). The correlation for RSV targets was maintained when broken down further by subtype (Supplemental Figure 1). Variance between the Ct values reported by each assay was also low for all targets, with an average Ct value standard deviation of 0.22 for influenza A, 0.24 for influenza B, and 0.36 for RSV (Supplemental Data 1). When RSV was analyzed by subtype, average Ct value standard deviation was greatest for RSVA at 0.72 (n=7), with measured Ct values an average of 1.4 cycles lower with the *PLUS* Combination Test (Supplemental Figure 1; Supplemental Data 1). The average run time from insertion of the cartridge to its ejection was timed for both the *PLUS* Combination Test and SARS-CoV-2/Flu/RSV Combination Test. There was no difference in run time between the two assays, with an average of 36.5 minutes to assay completion (Supplemental Data 1).

**Table 3:** Concordance of the Xpert SARS-CoV-2/Flu/RSV Combination Test with the SARS-CoV-2/Flu/RSV *PLUS* Combination Test for the detection of Influenza A, Influenza B and RSV.

| Result with<br>Xpert SARS-CoV-2/Flu/RSV | Result with Xpert SARS-CoV-2/Flu/RSV *PLUS* |  |  |  |  |  |
| --- | --- | --- | --- | --- | --- | --- |
|  | Influenza A |  | Influenza B |  | RSVA/B |  |
|  | Positive | Negative | Positive | Negative | Positive | Negative |
| Positive | 21 | 0 | 19 | 1 | 25 | 0 |
| Negative | 0 | 139 | 0 | 140 | 0 | 135 |

## Discussion and Conclusion

In an effort to improve assay performance against emerging SARS-CoV-2 variants the *PLUS* Combination Test introduces an additional SARS-CoV-2 target (RdRp), as this region has is unmodified in all circulating variants of concern (te Velthuis *et al*., 2009; Pachetti *et al*., 2020; Peng *et al*., 2020), and has introduced redesigned N2 oligos for better coverage of circulating mutations that caused N gene target dropout (Foster *et al*., 2022; Isabel *et al*., 2022). Here, we investigated the performance characteristics of the *PLUS* Combination Test comparing it to the SARS-CoV-2/Flu/RSV combination test authorized for use in Canada since January, 2021. This study used a total of 160 clinical samples (including mixtures) that were processed at two independent institutions, of which 110 samples were positive for SARS-CoV-2, influenza A, influenza B, RSVA, and/or RSVB. There was no observable difference in assay run time, sensitivity, specificity, or reported Ct value between the *PLUS* Combination Test and SARS-CoV-2/Flu/RSV combination test – with the possible exception of a slight decrease in RSVA Ct value with the *PLUS* Combination Test across a small number of samples. Overall, the results between the two assays were nearly identical for all targets, with agreement approaching 100%. The only discordant sample between the two assays was a high-Ct (32.3) Influenza B sample detected only with the SARS-CoV-2/Flu/RSV Combination Test. This likely reflects detection variability due to the low concentration of the target near its LOD, rather than a difference in performance between the two assays.

This observation was consistent for all variants tested. These results demonstrate that the Ct value for RdRp detection of SARS CoV-2 likely falls at or above the E and N2 targets producing little to no change to the reported Ct value. In conclusion, as the performance characteristics of the tests are nearly identical, the *PLUS* Combination Test can effectively replace the SARS-CoV-2/Flu/RSV combination test.

## Data Availability

All primary research data used to generate this manuscript are included in-text and within Supplementary file 1.

The authors have no competing interests to disclose.

All primary research data used to generate this manuscript are included in-text and within Supplementary file 1.

## References

Becker, M. G. et al. (2020) ‘Recommendations for sample pooling on the Cepheid GeneXpert® system using the Cepheid Xpert® Xpress SARS-CoV-2 assay’, PLOS ONE. Edited by J.-L. E. Darlix. Public Library of Science, 15(11), p. e0241959. doi: 10.1371/journal.pone.0241959.

Berry, L. et al. (2020) ‘Point of care testing of Influenza A/B and RSV in an adult respiratory assessment unit is associated with improvement in isolation practices and reduction in hospital length of stay’, Journal of Medical Microbiology, 69(5), pp. 697–704. doi: 10.1099/jmm.0.001187.

Broder, K. et al. (2020) ‘Test agreement between roche cobas 6800 and cepheid genexpert xpress sars-cov-2 assays at high cycle threshold ranges’, Journal of Clinical Microbiology, 58(8). doi: 10.1128/JCM.01187-20.

Cepheid Package Insert (2021) ‘Xpert ® Xpress SARS-CoV-2/Flu/RSV Instructions For Use with GeneXpert Dx or GeneXpert Infinity Systems’. Cepheid, 302–5707(Rev. A).

Foster, C. S. et al. (2022) ‘SARS-CoV-2 N-gene mutation leading to Xpert Xpress SARS-CoV-2 assay instability’, Pathology, 54(4): 499–501.

Goldenberger, D. et al. (2020) ‘Brief validation of the novel GeneXpert Xpress SARS-CoV-2 PCR assay’, Journal of Virological Methods, 284(July), pp. 8–10. doi: 10.1016/j.jviromet.2020.113925.

Gotham, D. et al. (2021) ‘Public investments in the development of GeneXpert molecular diagnostic technology’, PLoS ONE, 16(8 August). doi: 10.1371/journal.pone.0256883.

Hogan, C. A. et al. (2020) ‘Five-minute point-of-care testing for SARS-CoV-2: Not there yet’, Journal of Clinical Virology. Elsevier B.V., 128. doi: 10.1016/J.JCV.2020.104410.

Isabel, S. et al. (2022) ‘Emergence of a mutation in the nucleocapsid gene of SARS-CoV-2 interferes with PCR detection in Canada’, Scientific Reports, 12(1), 1–7.

Johnson, G. et al. (2021) ‘Clinical evaluation of the GeneXpert® Xpert® Xpress SARS-CoV-2/Flu/RSV combination test’, Journal of Clinical Virology Plus, 1(1), pp. 100014. doi: 10.1016/j.jcvp.2021.100014.

Johnson, G. et al. (2022) ‘ Clinical Evaluation of the GeneXpert® Xpert® Xpress SARS-CoV-2/Flu/RSV PLUS Combination Test’, medRxiv. Cold Spring Harbour Laboratory Press, p. 2022.11.07.22281957. doi: https://doi.org/10.1101/2022.11.07.22281957

Jokela, P. et al. (2020) ‘SARS-CoV-2 sample-to-answer nucleic acid testing in a tertiary care emergency department: evaluation and utility’, Journal of Clinical Virology. Elsevier B.V., 131. doi: 10.1016/J.JCV.2020.104614.

Lieberman, J. et al. (2020) ‘Comparison of Commercially Available and Laboratory Developed Assays for in vitro Detection of SARS-CoV-2 in Clinical Laboratories’, medRxiv. Cold Spring Harbor Laboratory Press, p. 2020.04.24.20074559. doi: 10.1101/2020.04.24.20074559.

Moran, A. et al. (2020) ‘The Detection of SARS-CoV-2 using the Cepheid Xpert Xpress SARS-CoV-2 and Roche cobas SARS-CoV-2 Assays’, Journal of Clinical Microbiology. American Society for Microbiology Journals. doi: 10.1128/JCM.00772-20.

Pachetti, M. et al. (2020) ‘Emerging SARS-CoV-2 mutation hot spots include a novel RNA-dependent-RNA polymerase variant’, J Transl Med, 18, p. 179. doi: 10.1186/s12967-020-02344-6.

Peng, Q. et al. (2020) ‘Structural and Biochemical Characterization of the nsp12-nsp7-nsp8 Core Polymerase Complex from SARS-CoV-2’, Cell Reports, 31(11). doi: 10.1016/j.celrep.2020.107774.

Rakotosamimanana, N. et al. (2020) ‘GeneXpert for the diagnosis of COVID-19 in LMICs’, The Lancet Global Health. Elsevier Ltd, 8(12), pp. e1457–e1458. doi: 10.1016/S2214-109X(20)30428-9.

Respiratory Virus Infections Working Group (2020) ‘Canadian Public Health Laboratory Network: Prioritized support for northern, remote and isolated communities in Canada’, Can Commun Dis Rep, 46(10), pp. 322–323. doi: 10.14745/ccdr.v46i10a02.

Rong, K. et al. (2022) ‘Validation of the Cepheid Xpert® Xpress SARS-CoV-2 using upper and lower respiratory tract specimens’, European Journal of Microbiology and Immunology. Akadémiai Kiadó, 12(1), pp. 18–21. doi: 10.1556/1886.2022.00003.

Smithgall, M. C. et al. (2020) ‘Comparison of Cepheid Xpert Xpress and Abbott ID Now to Roche cobas for the Rapid Detection of SARS-CoV-2’, bioRxiv. Cold Spring Harbor Laboratory, p. 2020.04.22.055327. doi: 10.1101/2020.04.22.055327.

Tham, J. W. M. et al. (2021) ‘Parallel testing of 241 clinical nasopharyngeal swabs for the detection of SARS-CoV-2 virus on the Cepheid Xpert Xpress SARS-CoV-2 and the Roche cobas SARS-CoV-2 assays’, Clinical Chemistry and Laboratory Medicine. De Gruyter Open Ltd, 59(2), pp. E45–E48. doi: 10.1515/CCLM-2020-1338/MACHINEREADABLECITATION/RIS.

te Velthuis, A. J. W. et al. (2009) ‘The RNA polymerase activity of SARS-coronavirus nsp12 is primer dependent’, Nucleic Acids Research, 38(1), pp. 203–214. doi: 10.1093/nar/gkp904.

Wolters, F. et al. (2020) ‘Multi-center evaluation of cepheid xpert® xpress SARS-CoV-2 point-of-care test during the SARS-CoV-2 pandemic’, Journal of Clinical Virology. Elsevier, 128(May 2020), p. 104426. doi: 10.1016/j.jcv.2020.104426.

World Health Organization (2020) ‘Rapid communication on the role of the GeneXpert ® platform for rapid molecular testing for SARS-CoV-2 in the WHO European Region European Laboratory Initiative on TB, HIV and Viral Hepatitis: 1 April 2020’, (April). Available at: https://apps.who.int/iris/handle/10665/336322 (Accessed: 9 September 2022).

Yau, F. et al. (2021) ‘Clinical utility of a rapid “on-demand” laboratory-based SARS-CoV-2 diagnostic testing service in an acute hospital setting admitting COVID-19 patients’, Clinical Infection in Practice. Elsevier B.V., 12. doi: 10.1016/J.CLINPR.2021.100086.

Zhen, W. et al. (2020) ‘Clinical Evaluation of Three Sample-To-Answer Platforms for the Detection of SARS-CoV-2’, Journal of Clinical Microbiology. American Society for Microbiology Journals. doi: 10.1128/JCM.00783-20.

