## Supplemental Figure 1 for "Clinical Evaluation of the GeneXpert^®^ Xpert^®^ Xpress SARS-CoV-2/Flu/RSV *PLUS* Combination Test"

### Slide 1
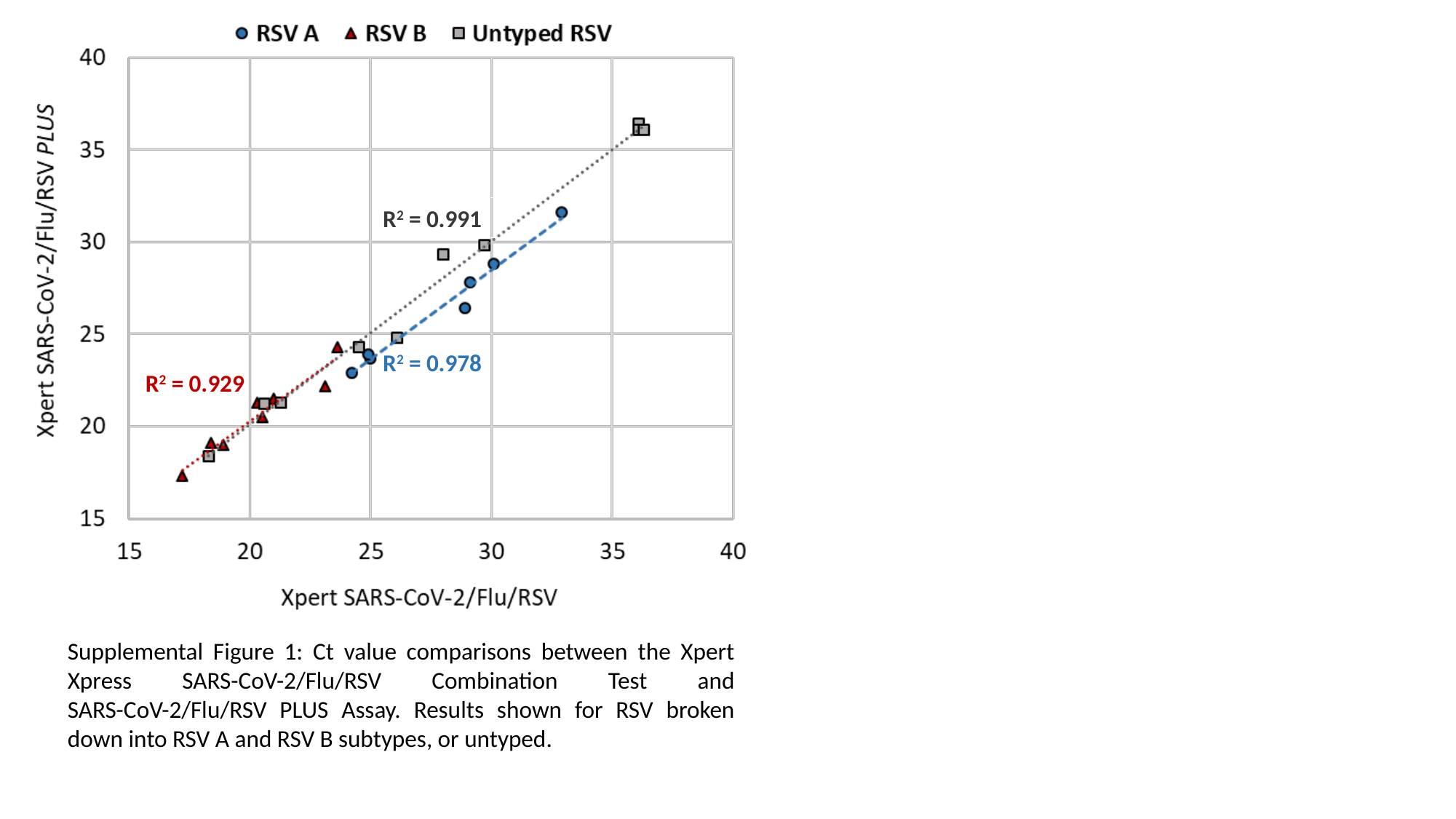

R^2^ = 0.991
R^2^ = 0.978
R^2^ = 0.929
Supplemental Figure 1: Ct value comparisons between the Xpert Xpress SARS-CoV-2/Flu/RSV Combination Test and SARS-CoV-2/Flu/RSV PLUS Assay. Results shown for RSV broken down into RSV A and RSV B subtypes, or untyped.
